# Residual Radiation Risk Disparities Across Sex and Race or Ethnic Groups for Lifetime Never-Smokers on Lunar Missions

**DOI:** 10.1101/2023.06.04.23290952

**Authors:** Francis A. Cucinotta, Premkumar B. Saganti

## Abstract

In the current decade missions to the Earth’s moon are of scientific and societal interest, however pose the problem of risks of late effects for returning crew persons, most importantly cancer and circulatory diseases. In this paper, we discuss NSCR-2022 model risk estimates for lunar missions for US racial and ethnic groups comparing never-smokers to US averages for each group and sex. We show that differences within groups between men and women are largely reduced for NS compared to the average population. Race and ethnic group dependent cancer and circulatory disease risks are reduced by 10% to 40% for NS with the largest decrease for Whites. Circulatory disease risks are changed by less than 10% for NS and in several cases modestly increased due to increased lifespan for NS. Asian-Pacific Islander (API) and Hispanics NS are at lower risk compared to Whites and Blacks. Differences between groups is narrowed for NS compared to predictions for average populations, however disparities remain especially for Blacks and to a lesser extent Whites compared to API or Hispanic NS groups.

## Introduction

In the 1980’s the small population of specialized workers comprised of astronauts led the National Council on Radiation Protection and Measurements (NCRP) to recommend [1] an individualized risk assessment approach for radiation protection at NASA; essentially sex and age specific dose limits corresponding to a lifetime 3% fatality risk limit. This approach is similar to recommendations by the US National Institutes of Health (NIH) that maintains a policy that sex as a biological variable (SABV) is of utmost importance in clinical care and research [2]. The astronaut population has become more diverse and there is interest in privately funded spaceflights which suggests an increased focus on individualized based risk assessment is needed. At this time genetic screening-based approaches to risk limits are not possible due to legal limitations under the GINA laws [3], and importantly the scientific basis for such an approach for exposures to low doses or high LET radiation has not been developed or verified.

Beyond sex and age risk dependencies, the use of tobacco products is well known as the largest contributor to risks of cancer and other diseases such as circulatory diseases and chronic obstructive pulmonary disease (COPD) [4–6]. Indeed, other risk factors such as use of alcohol, obesity, and poor nutrition have reduced impact compared to the use of tobacco products, and are difficult to quantify. Because astronauts are predominantly lifetime NS [7] previous research by one of the authors [8,9] developed an approach to estimate space radiation risks for NS. Radiation risks are most often calculated within a multiplicative risk model using data for a general population, which inherently includes the impact of use of tobacco products by current and former smokers on risk estimates for never-smokers (NS). Recent estimates show that the proportion of smokers in the USA has declined to 10.1% and 13.1% for women and men, respectively in 2021 [10] from 18% and 24%, respectively in 2010 as used in an earlier report [8]. In addition, different proportions of smokers (S), former smokers (FS) and NS occur in various racial or ethnic groups, which impact population data and estimates of NS risks that are more representative of astronauts. Furthermore, there are also important racial and ethnic group disparities on cancer risks that were estimated in a recent report [11].

In this paper we extend these results to consider estimates of relative risks and population proportions of NS in the US categorized into several groups. This allows for sex and age specific risk estimates for NS in US populations of Asian-Pacific Islander (API), Black, Hispanic, and White groups. We apply the model to risk predictions for extended stays on the Earth’s moon near solar minimum conditions. As described previously [8,9,11] there are competing factors in estimates for NS and racial/ethnic groups, which are the influence of possible higher (or lower) background cancer rates in the multiplicative risk model for specific disease types versus a longer (or shorter) lifespan for a model population that possibly increases (or decreases) lifetime risk estimates. The prime example described in our earlier report [11] was comparison of risks for Blacks versus White populations where the influence of higher cancer rates in Blacks is reduced because of decreased lifespan leading to similar space radiation risk estimates. However, more favorable competing factors led to ∼30% lower estimates for Hispanics and Asian-Pacific Islanders (API) compared to Blacks or White populations.

Uncertainty analysis in radiation risk predictions are of importance because of the sparsity of epidemiology [12] and radiobiology data, especially for high linear energy transfer (LET) radiation [13,14]. A risk prediction is not scientifically valid without an assessment of the uncertainty in the prediction. The use of an effective dose maybe acceptable for low risk levels well below an acceptable risk limit, however as a limit is approached its use is confounded by large uncertainties thus requiring their evaluation. In addition, effective dose has reduced value in individualized risk assessment approaches. The NASA Space Cancer Risk (NSCR) model has evolved for nearly 25 years to consider changes in epidemiology data and new results from space radiobiology research [9,11,15-19]. In this paper the current version NSCR-2022 [11,18,19] is used to make predictions for extended lunar stays with application to sex and racial-ethnic group specific risk estimates for extended lunar stay missions. NSCR-2022 uses a radiation quality descriptor with estimates of the effects of non-targeted effects [18,19]. We consider scenarios of 35-year old astronauts on 60-day lunar stays and 45-year old astronauts with 180-day lunar stays, while assuming missions occur at an average solar minimum where galactic cosmic ray (GCR) exposures are near their highest values.

## Methods

We briefly summarize the method developed to predict the risk of exposure induced cancer (REIC) and risk of exposure induced death (REID) for space missions and associated uncertainty distributions [17,18]. The instantaneous cancer incidence or mortality rates, λ_I_ and λ_M_, respectively, are modeled as functions of the tissue averaged absorbed dose *D_T_*, or dose-rate *D_Tr_*, gender, age at exposure *a_E_*, and attained age *a* or latency *L,* which is the time after exposure *L=a-a_E_*. The λ_I_ (or λ_M_) is a sum over rates for each tissue that contributes to cancer risk, λ_IT_ (or λ_MT_). These dependencies vary for each cancer type that could be increased by radiation exposure. The REIC is calculated by folding the instantaneous radiation cancer incidence-rate with the probability of surviving to time *t,* which is given by the survival function *S_0_(t)* for the background population times the probability for radiation cancer death at previous time, summing over one or more space mission exposures, and then integrating over the remainder of a lifetime:

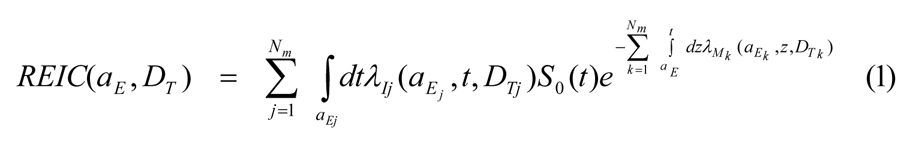

where z is the dummy integration variable. In equation (1), N_m_ is the number of missions (exposures), and for each exposure, j, there is a minimum latency of 5-years for solid cancers and circulatory diseases, and 2-years for leukemia assumed. Tissue specific REIC estimates are similar to Eq. (1) using the single term from λ_I_ of interest. The equation for the REID estimate is also similar to Eq. (1) with the incidence rate replaced by the mortality rate (defined below). We terminate the integral in Eq.(1) at the age of 100 years.

The tissue-specific cancer incidence rate for an organ absorbed dose, *D_T_*, is written as a weighted average of the multiplicative and additive transfer models, denoted as a mixture model. However, a scaling factor, *R_QF_* is introduced for extrapolating to low dose and dose-rates and estimating the radiation quality dependences of cancer risk for a particle of charge number Z and kinetic energy per nucleon, E:

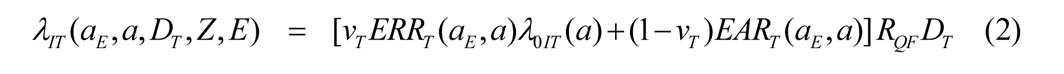

where *v_T_* is the tissue-specific transfer model weight, *λ_0IT_* is the tissue-specific cancer incidence rate in the reference population, and where *ERR_T_* and *EAR_T_* are the tissue specific excess relative risk and excess additive risk per Sievert, respectively. Extension of Eq. (2) for a spectrum of particles was described previously [17–19].

The tissue specific rates for cancer mortality *λ_MT_* are modeled following the BEIR VII report [20] whereby the incidence rate of Eq. (2) is scaled by the age, sex, and tissue specific ratio of rates for mortality to background incidence in the population under study:

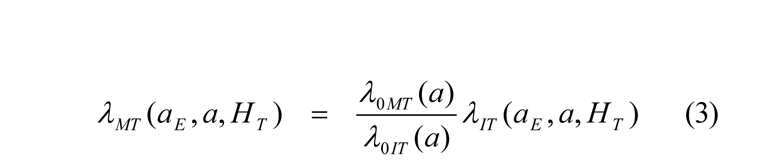

Lifetables from the U.S. Center of Disease Control and Prevention (CDC) for Black, White, and Hispanic (Black and White) male and females are used [21], while the life-table for Asian-Pacific Islander populations are adjusted from the data for the Hispanics for their higher life expectance of ∼2 years. Total and tissue specific cancer incidence and mortality rates from U.S. SEER are used [22], which provide race, age, and sex specific rates. For cancer incidence we used the SEER delay-adjusted rates, which estimate the delay in reporting of cancer cases [23]. In several cases (e.g., liver and brain cancers), the delay-adjusted rates led at older ages (>80 y) to age-specific mortality rates greater than the delay adjusted incidence rate. For these cases we applied a correction to mortality rates to ensure the mortality rate was 10% larger than the delay adjusted age specific incidence rates. Data for non-cancer risks (COPD, ischemic heart disease (IHD), cerebrovascular disease and other heart disease) are from the CDC data base [24].

We use ERR and EAR functions for tissue specific cancer incidence from the most recent series of reports from the Radiation Effects Research Foundation (RERF) [25–32] on the Life Span Study of Atomic-Bomb survivors (LSS). This RERF series has improved considerations of lifestyle factor such as use of tobacco products and alcohol compared to earlier reports, and uses the more recent LSS dosimetry system. The BEIR VII report recommended a transfer weight of 0.7 for the ERR model (0.3 for the EAR model), which we apply except for a few cases. For lung cancer we use the generalized ERR model [25], which accounts for cigarette usage explicitly. EAR functions from Preston et al [33] were used for the remainder term, while no EAR functions were found for several cancers with smaller numbers of cases in the Life Span Study (LSS) of atomic bomb survivors. Here we use only the ERR model for esophagus, oral cavity, brain, prostate, ovarian and pancreatic cancers. For circulatory risk estimates we use the recent meta-analysis results for ERR estimates [34].

### Never-Smoker Risk Factors

We use CDC estimates of proportions of smokers (S) and former smokers (FS) in the U.S. population [35,36], which are show in **Table 1**. Relative risks for S, FS and NS for tissue specific cancers, circulatory diseases and COPD mortality are from the meta-analysis of Carter et al. [6]. We use these values for both cancer incidence and mortality predictions. To implement these results, we consider cancer rates reported for the US race or ethic specific population groups comprised of populations of S, FS, and NS with proportions *f_S_*, *f_FS_*, and *f_NS_*,

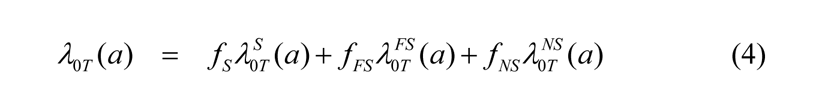

The relative risks (RR) of S and FS compared to NS, *RR_S_* and *RR_FS_*, respectively are then used to compare rates for NS to the U.S. average rates for each group:

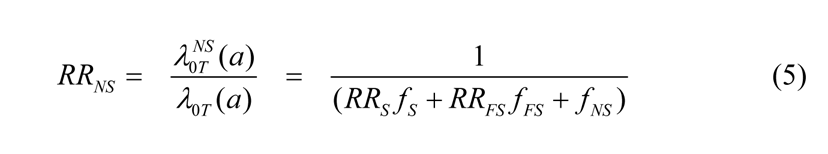

**Table 1.** Percentage of US populations by race or ethnic group of current smokers, former smokers and lifetime never-smokers [35,36].

| **Race/Ethnic Group** | **% Current Smokers** | **%Former Smokers** | **%Never-Smokers** |
| --- | --- | --- | --- |
|  | **Females** |  |  |
| **API** | 2.6 | 4.0 | 93.4 |
| **Black** | 13.5 | 10 | 76.5 |
| **Hispanic** | 7.1 | 8.4 | 84.5 |
| **White** | 16.0 | 19.7 | 64.3 |
|  | **Males** |  |  |
| **API** | 12.0 | 14.0 | 74.0 |
| **Black** | 20.9 | 15.7 | 63.4 |
| **Hispanic** | 13.1 | 15.6 | 71.3 |
| **White** | 17.3 | 24.4 | 58.3 |

The resulting estimates of *RR_NS_* are shown in **Tables 2** for females and **Tables 3** for males. Possible race or ethnic group dependencies in *RR_S_*, *RR_FS_* and *RR_NS_* are discussed below. For Monte-Carlo uncertainty evaluations we assume normal distribution with standard deviations as reported [6].

**Table 2.** Relative risks (RR) of tissue specific cancers, ischemic heart disease (IHD), cerebrovascular disease and chronic obstructive pulmonary disease (COPD) for US Females of different races or ethnic groups. Relative risks for current (S) and former smokers (FS) against never-smoker (NS) baseline [6] are used to estimate RR for NS in US population data as described in the text.

| **FEMALES** | **RR(S)** | **RR(FS)** | **RR(NS)** | **RR**<br>**(NS to Ave)** | **RR**<br>**(NS to Ave)** | **RR**<br>**(NS to Ave)** | **RR**<br>**(NS to Ave)** |
| --- | --- | --- | --- | --- | --- | --- | --- |
| **RISK:** |  |  |  | **White** | **Black** | **Asian** | **Hispanic** |
| **Leukemia** | 1.1 | 1.1 | 1 | 0.97 | 0.98 | 0.99 | 0.98 |
| **Stomach** | 1.7 | 1.1 | 1 | 0.88 | 0.91 | 0.98 | 0.95 |
| **Colon** | 1.6 | 1.2 | 1 | 0.88 | 0.91 | 0.98 | 0.94 |
| **Liver** | 1.8 | 1.1 | 1 | 0.87 | 0.89 | 0.98 | 0.94 |
| **Bladder** | 3.9 | 2.3 | 1 | 0.58 | 0.66 | 0.89 | 0.76 |
| **Lung** | 22.9 | 6.8 | 1 | 0.18 | 0.22 | 0.56 | 0.33 |
| **Esophagus** | 6.4 | 5.1 | 1 | 0.37 | 0.47 | 0.77 | 0.58 |
| **Oral Cavity** | 5.6 | 2.2 | 1 | 0.51 | 0.57 | 0.86 | 0.70 |
| **Brain** | 1 | 1 | 1 | 1.00 | 1.00 | 1.00 | 1.00 |
| **Thyroid** | 1 | 1 | 1 | 1.00 | 1.00 | 1.00 | 1.00 |
| **Pancreas** | 1.9 | 1.2 | 1 | 0.84 | 0.88 | 0.97 | 0.93 |
| **Remainder** | 2.7 | 1.4 | 1 | 0.74 | 0.79 | 0.94 | 0.87 |
| **Breast** | 1.3 | 1.2 | 1 | 0.92 | 0.94 | 0.98 | 0.96 |
| **Ovarian** | 1 | 1 | 1 | 1.00 | 1.00 | 1.00 | 1.00 |
| **COPD** | 25 | 9.2 | 1 | 0.15 | 0.20 | 0.51 | 0.29 |
| **IHD** | 3 | 1.6 | 1 | 0.70 | 0.75 | 0.93 | 0.84 |
| **Cerebrovascular** | 2.1 | 1.2 | 1 | 0.82 | 0.86 | 0.96 | 0.91 |
| **Other Heart Disease** | 1.9 | 1.4 | 1 | 0.82 | 0.86 | 0.96 | 0.91 |

**Table 3.** Relative risks (RR) of tissue specific cancer, ischemic heart disease (IHD), cerebrovascular disease and chronic obstructive pulmonary disease (COPD) for US Males of different races or ethnic groups. Relative risks for current (S) and former smokers (NS) against never-smoker (NS) baseline [6] are used to estimate disease specific RR for NS from US population data as described in the text.

| **MALES** | **RR(S)** | **RR(FS)** | **RR(NS)** | **RR**<br>**(NS to Ave)** | **RR**<br>**(NS to Ave)** | **RR**<br>**(NS to Ave)** | **RR**<br>**(NS to Ave)** |
| --- | --- | --- | --- | --- | --- | --- | --- |
| **Risk:** |  |  |  | **White** | **Black** | **Asian** | **Hispanic** |
| **Leukemia** | 1.9 | 1.4 | 1 | 0.80 | 0.80 | 0.86 | 0.84 |
| **Stomach** | 1.9 | 1.5 | 1 | 0.78 | 0.79 | 0.85 | 0.84 |
| **Colon** | 1.4 | 1.2 | 1 | 0.89 | 0.90 | 0.93 | 0.92 |
| **Liver** | 2.3 | 1.5 | 1 | 0.74 | 0.74 | 0.82 | 0.80 |
| **Bladder** | 3.27 | 2.09 | 1 | 0.60 | 0.61 | 0.70 | 0.68 |
| **Lung** | 25.3 | 6.8 | 1 | 0.15 | 0.14 | 0.21 | 0.20 |
| **Esophagus** | 3.9 | 2.6 | 1 | 0.53 | 0.54 | 0.64 | 0.61 |
| **Oral Cavity** | 5.7 | 1.7 | 1 | 0.50 | 0.48 | 0.60 | 0.58 |
| **Brain** | 1 | 1 | 1 | 1.00 | 1.00 | 1.00 | 1.00 |
| **Thyroid** | 1 | 1 | 1 | 1.00 | 1.00 | 1.00 | 1.00 |
| **Pancreas** | 1.6 | 1 | 1 | 0.91 | 0.89 | 0.93 | 0.93 |
| **Remainder** | 3.2 | 1.5 | 1 | 0.67 | 0.65 | 0.75 | 0.73 |
| **Breast** | 1 | 1 | 1 | 1.00 | 1.00 | 1.00 | 1.00 |
| **Prostate** | 1.4 | 1 | 1 | 0.94 | 0.92 | 0.95 | 0.95 |
| **COPD** | 27.8 | 7.5 | 1 | 0.14 | 0.13 | 0.20 | 0.18 |
| **IHD** | 2.6 | 1.5 | 1 | 0.72 | 0.71 | 0.79 | 0.78 |
| **Cerebrovascular** | 1.9 | 1.2 | 1 | 0.83 | 0.82 | 0.88 | 0.87 |
| **Other Heart Disease** | 2 | 1.3 | 1 | 0.80 | 0.80 | 0.86 | 0.85 |

### Space Radiation Quality Descriptor

The *R_QF_* is estimated from relative biological effectiveness factors (RBE’s) and dose-rate modifiers determined from low dose and dose-rate particle data relative to acute γ-ray exposures for doses of about 0.5–3 Gy, which we denote as *RBE_γAcute_* to distinguish from *RBE_max_*, which is based on less accurate initial slope estimates for γ-rays. The quality function, *R_QF_*, considers several contributions. The first representing an ion track’s penumbra of energetic δ-rays (electrons), which are assumed to follow the dose response estimated from γ-ray epidemiology studies. Here since we are scaling directly to the acute γ-ray responses, the dose-rate modifier is assumed to be influenced by results from experimental models used in RBE determinations. The dose-rate modifier (assumed to be similar to estimate of a dose and dose-rate reduction effectiveness factor (DDREF)) adjusts the penumbra like term. The second term represents the ions track core of ultra-high ionizations whereby no dose-rate modifier is assumed. Radiobiological experiments that vary the charge number and velocity of an ion are optimal to determine the relative contributions of the penumbra and core components. The final term represents the contribution of non-targeted effects (NTE) as described below.

In this approach the scaling factor for the penumbra-like and core-like terms of low and high ionization densities, respectively is based on the following parametric function with parameters fit to experimental data [17–19] denoted as the targeted effects (TE) terms:

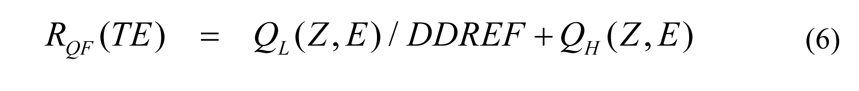

where

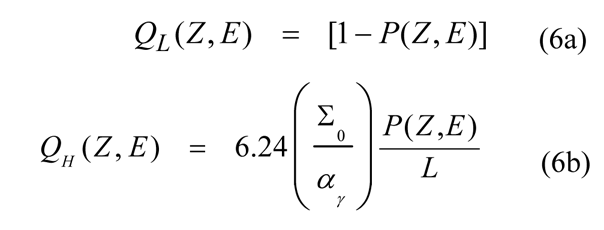

with the parametric function [17]

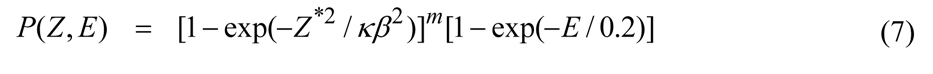

where *E* is the particles kinetic energy per nucleon, *L* is the LET in keV/μm, *Z* is the particles charge number, *Z\** the effective charge number, and *β* the particles speed relative the speed of light. An ancillary condition is used to correlate the values of the parameter κ as a function of *m* as [17]

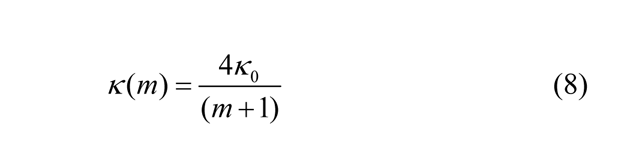

where κ_0_ is value for the most likely value *m*=3. This constraint fixes the peak effectiveness with kinetic energy for each heavy ion charge group in the model to be consistent with results from experiments [17]. The three model parameters (Σ_0_/α_γ_, κ and *m*) in Eq. (6) are fit to radiobiology data for tumors in mice or surrogate cancer endpoints as described previously [17–19] as shown in **Table 4**. Distinct parameters are used for estimating solid cancer and leukemia risks based on estimates of smaller RBEs for acute myeloid leukemia and thymic lymphoma in mice compared to those found for solid cancers [17].

**Table 4.** Space Radiation Quality Factor Model Parameters with standard deviation (SD) in parameter estimate*.

| **Model**<br>**Parameter** | **High Z (Z>4)** | **Low Z (Z≤4)** |
| --- | --- | --- |
| **Slope**<br>**parameter, m** | 3±0.5 | 3±0.5 |
| **κ** | 624±69 | 1000±150 |
| **Solid Cancer** |  |  |
| **Σ<sub>0</sub>/α<sub>γ</sub>, μm<sup>2</sup>Gy<sup>-1</sup>** | (2897±357)/6.24 | (2897±250)/6.24 |
| **Leukemia** |  |  |
| **Σ<sub>0</sub>/α<sub>γ</sub>, μm<sup>2</sup>Gy<sup>-1</sup>** | (1750±250)/6.24 | (1750±250)/6.24 |
| **Parameters for NTE model** |  |  |
| **η<sub>0</sub>/α<sub>γ</sub>, Gy<sup>-1</sup>** | (6±3)x10<sup>-5</sup> | (6±3)x10<sup>-5</sup> |
| **η<sub>1</sub>** | 833±200 | 1000±250 |
| **A<sub>bys</sub>, μm<sup>2</sup>** | 1000±333 | 1000±333 |

\*Values of Σ<sub>0</sub>/α<sub>γ</sub> for solid cancer are shown without inclusion of mouse data for hepatocellular carcinoma and Harderian gland tumors whose inclusion leads to a higher value of (4728±1378)/6.24 as described previously [17].

In the NTE model we assume the TE contribution described above is valid with a linear response to the lowest dose or fluence considered, while an additional NTE contribution occurs [17–19]:

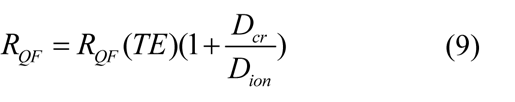

where *D_cr_* is found as the dose (or fluence) where the NTE and TE are equivalent that is given by

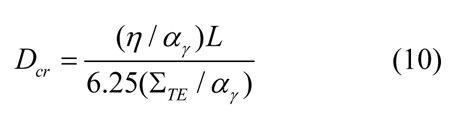

In terms of *F* the particle fluence (in units of 1/µm^2^), the η function represents the NTE contribution, which is parameterized as a function of *x=Z^*2^/β^2^* as:

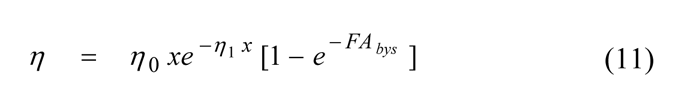

In Eq. (11) the area, *A_bys_*, reflects the region of bystander cells surrounding a cell traversed directly by a particle that receives an oncogenic signal.

Calculations are made using the HZETRN code and models of the GCR environments and radiation transport in spacecraft materials and tissue, which estimate the particle energy spectra, *ϕ_j_(E)* for 190 isotopes of the elements from Z=1 to 28, neutrons, and contributions from pions as described previously [17,19,37,38]. Organ doses from GCR and albedo particles are described in previous reports.

## Results

**Figures 1 to 4** show our modeled life-tables for NS and average groups resulting from the estimates in **Tables 1 to 3**. There is an increase in lifespan for NS of each group, however the disparity for Blacks and to a lesser extent Whites compared to API and Hispanics is apparent. In contrast for API and Hispanics the results for NS is close to that of the Average groups.

**Figure 1.**
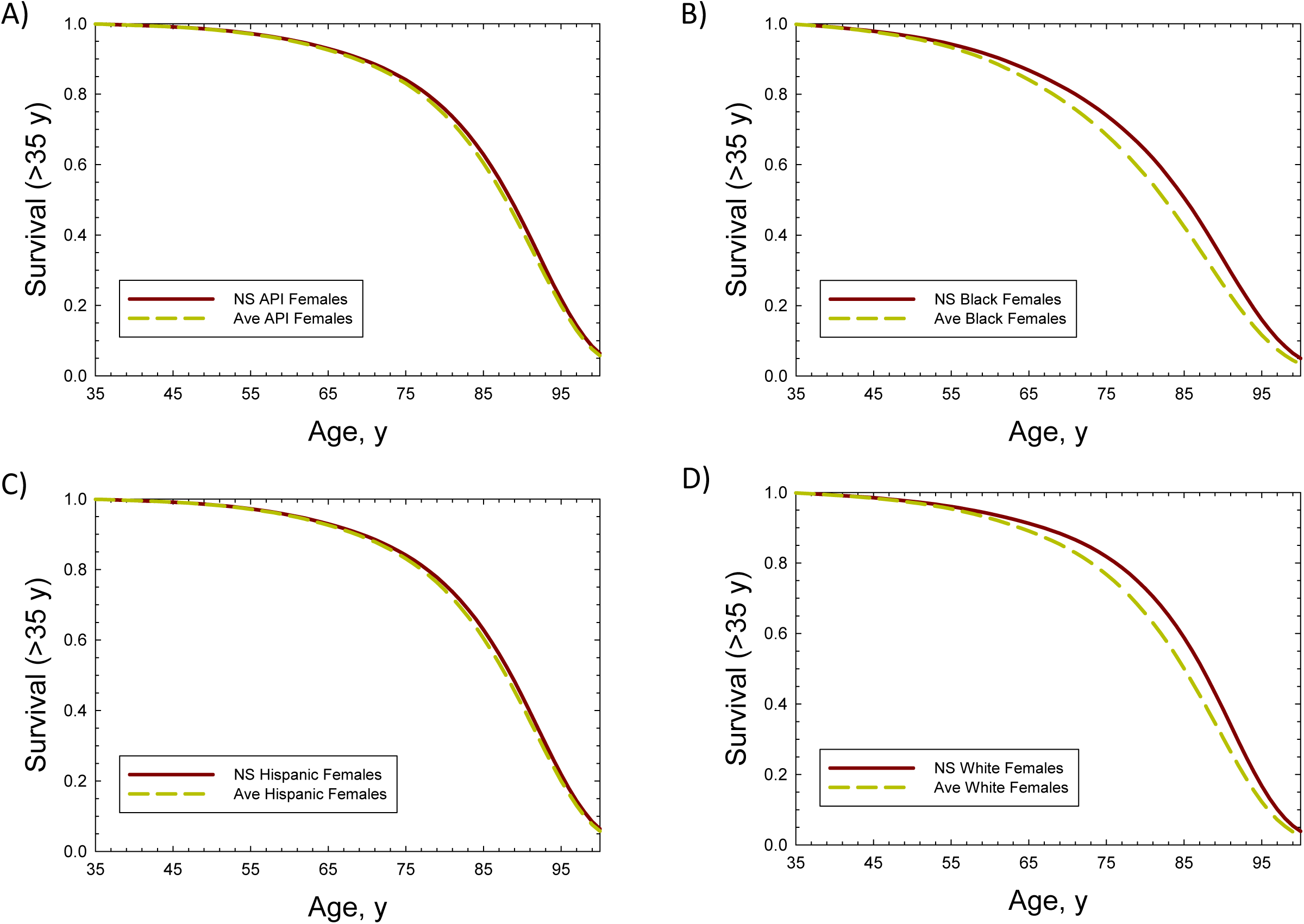
Conditional survival probabilities for Females with entry at age 35-y versus age comparing US average and Never-Smokers (NS) for A) Asian-Pacific Islanders, B) Black, C) Hispanic and D) Whites.

**Figure 2.**
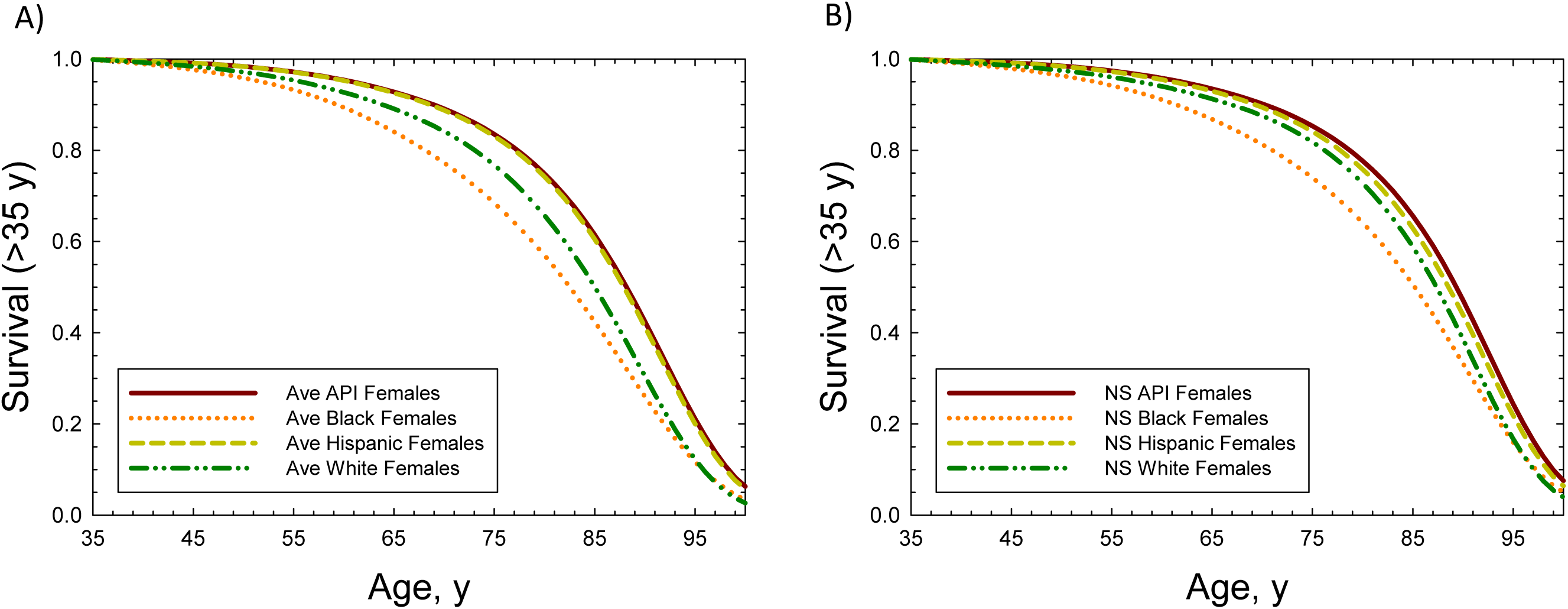
Conditional survival probabilities for Females with entry at age 35-y versus age comparing US average and Never-Smokers (NS) for different racial or ethnic groups. A) Average population, B) Never-smokers.

**Figure 3.**
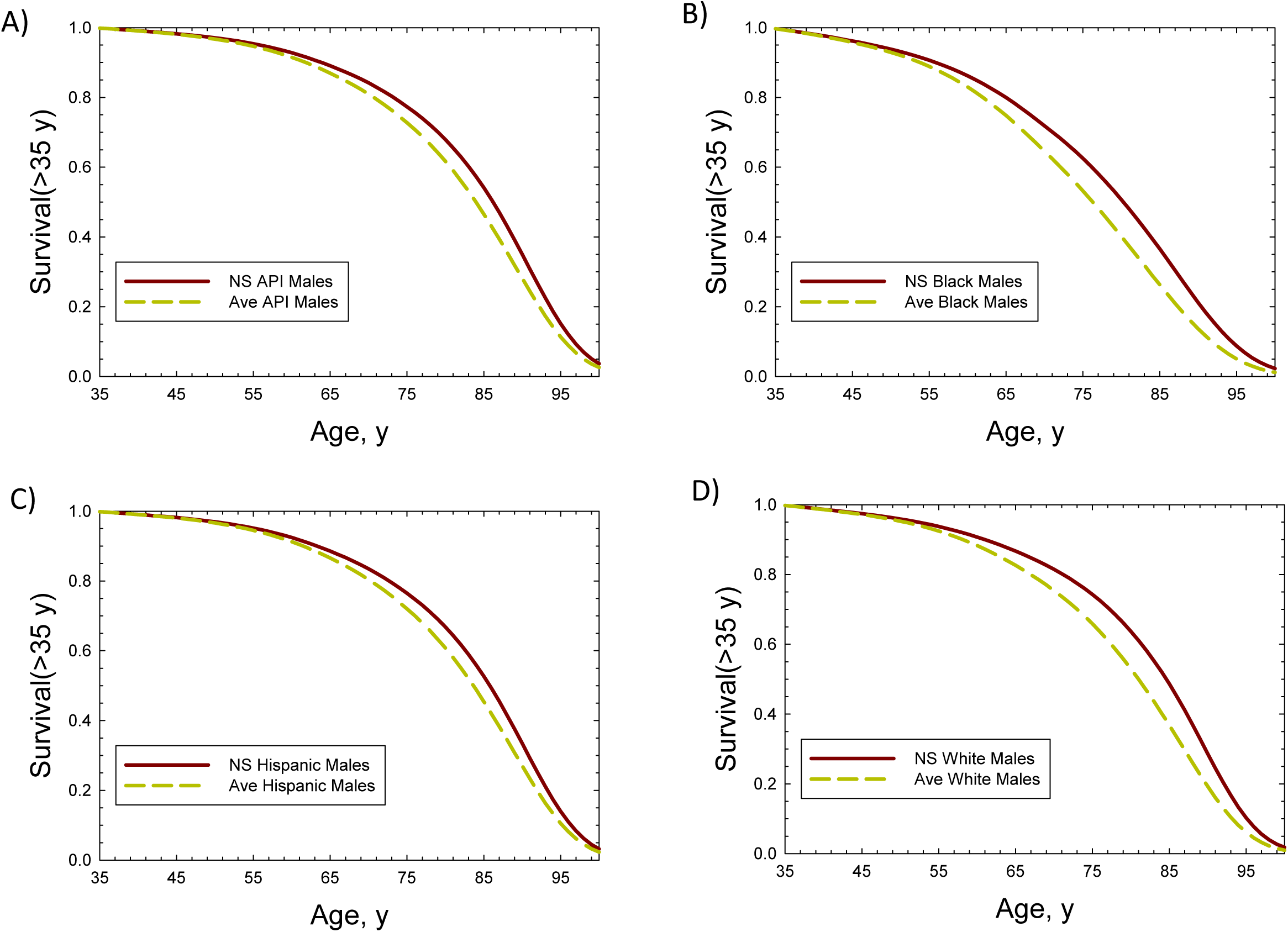
Conditional survival probabilities for Males with entry at age 35-y versus age comparing US average and Never-Smokers (NS) for A) Asian-Pacific Islanders, B) Black, C) Hispanic and D) Whites.

**Figure 4.**
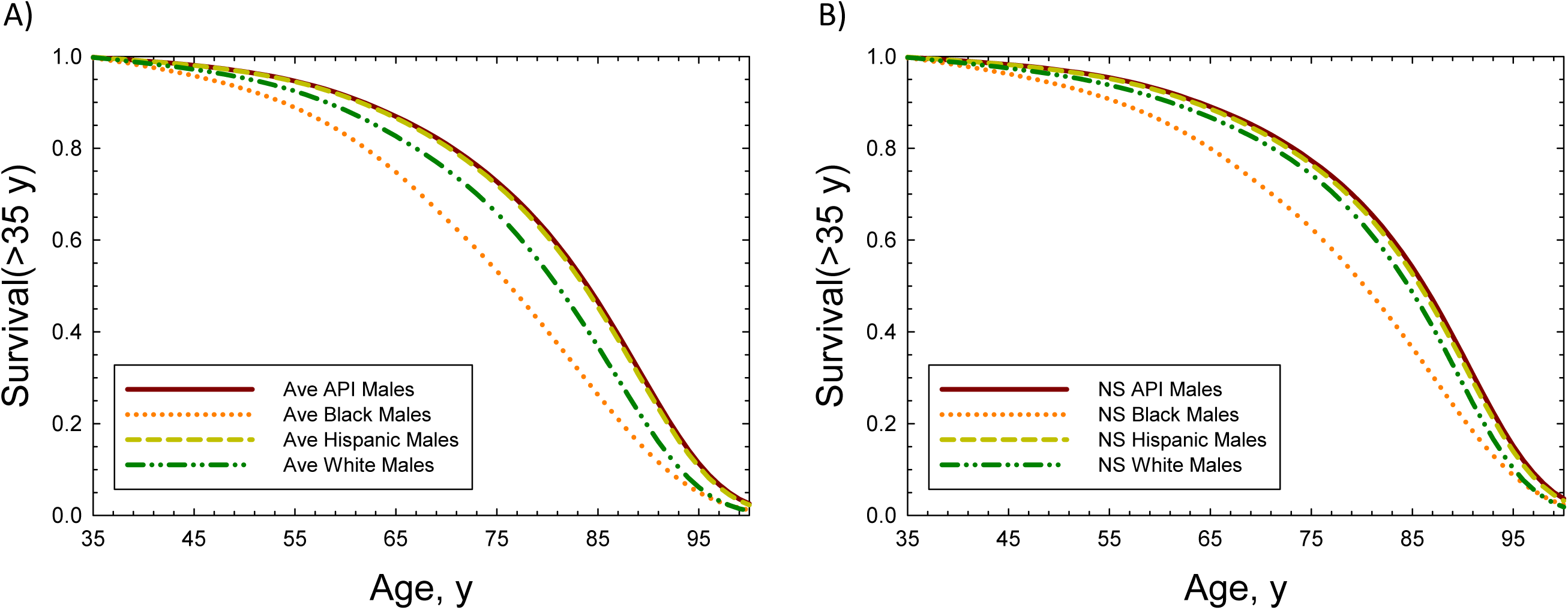
Conditional survival probabilities for Males with entry at age 35-y versus age comparing US average and Never-Smokers (NS) for different racial or ethnic groups. A) Average population, B) Never-smokers.

We considered risk predictions for 35-year old astronauts on lunar missions with 60 days on the lunar surface and 20 days total transit time between the Earth and moon [39]. Solar minimum conditions with 20 g/cm^2^ average aluminum shielding are used in the calculations. Predictions are shown in **Table 5** for females and **Table 6** for males. White females have the highest cancer risk for both average and NS groups. The inclusion of circulatory risks reduces differences for NS between API, Hispanics and Whites, however Blacks have a total REID about 20% larger than the API group. For males, Whites have the largest risks for the Average groups and group differences are reduced for NS compared to Average groups, however Black and White NS are ∼30% and ∼25% larger than the API NS total REID predictions. Sex dependent differences are also reduced between groups for NS with reductions from 20% to 10% and noting the inclusion of circulatory risks in total REID values reduces sex differences due to that from cancer risk alone.

**Table 5.** Cancer and circulatory disease risks and 95% confidence intervals for 35-year old Female astronauts of several race or ethnic groups comparing never-smokers to average groups population in the USA. Lunar missions near solar minimum with 20 g/cm^2^ aluminum shield is considered in calculations. Mission total duration is 80 days with 20 days for Earth to Moon transit and return and 60 days on lunar surface.

| **Race-Ethnic Group** | **%REIC** | **%REID-cancer** | **%REID-circulatory** | **%REID-Total** |
| --- | --- | --- | --- | --- |
|  | **Average Population** |  |  |  |
| **API** | 2.14 [0.72,5.3] | 0.72 [0.26,5.34] | 0.13 [0.05,0.26] | 0.86 [0.38,1.9] |
| **Blacks** | 2.24 [0.77,5.56] | 0.94 [0.33,2.24] | 0.18 [0.07,3.58] | 1.13 [0.49,2.45] |
| **Hispanic** | 2.15 [0.75,5.3] | 0.70 [0.25,1.65] | 0.13 [0.05,0.26] | 0.84 [0.37,1.8] |
| **Whites** | 2.72 [0.89, 6.84] | 1.01 [0.34,2.42] | 0.15 [0.06,0.28] | 1.12 [0.48,2.58] |
|  | **Never-Smokers** |  |  |  |
| **API** | 1.80 [0.59,4.54] | 0.56 [0.2,1.35] | 0.14 [0.05,0.28] | 0.71 [0.32,1.51] |
| **Blacks** | 1.78 [0.57,4.59] | 0.67 [0.23,1.61] | 0.21 [0.08,0.41] | 0.88 [0.39,1.83] |
| **Hispanic** | 2.01 [0.66,5.09] | 0.64 [0.22,1.53] | 0.14 [0.05,0.27] | 0.77 [0.33,1.67] |
| **Whites** | 2.00 [0.62,5.29] | 0.62 [0.20,1.53] | 0.17 [0.06,0.33] | 0.79 [0.35,1.71] |

**Table 6.** Cancer and circulatory disease risks and 95% confidence intervals for 35-year old Male astronauts of several race or ethnic groups comparing Never-Smokers to average group population in the USA. Lunar missions near solar minimum with 20 g/cm^2^ aluminum shield is considered in calculations. Mission total duration is 80 days with 20 days for Earth to Moon transit and return and 60 days on lunar surface.

| **Race-Ethnic Group** | **%REIC** | **%REID-cancer** | **%REID-circulatory** | **%REID-Total** |
| --- | --- | --- | --- | --- |
|  | **Average Population** |  |  |  |
| **API** | 1.48 [0.56,3.42] | 0.55 [0.21,1.25] | 0.15 [0.06,0.29] | 0.70 [0.33,1.42] |
| **Blacks** | 1.91 [0.71,4.45] | 0.74 [0.28,1.69] | 0.21 [0.08,0.42] | 0.96 [0.46,1.94] |
| **Hispanic** | 1.69 [0.63,3.89] | 0.61 [0.23,1.42] | 0.15 [0.06,0.42] | 0.77 [0.36,1.58] |
| **Whites** | 1.96 [0.74,4.54] | 0.74 [0.28,1.70] | 0.18 [0.07,0.36] | 0.93 [0.43,1.90] |
|  | **Never-Smokers** |  |  |  |
| **API** | 1.29 [0.47,3.04] | 0.46 [0.16,1.08] | 0.17 [0.06,0.32] | 0.62 [0.3,1.26] |
| **Blacks** | 1.71 [0.59,4.10] | 0.62 [0.22,1.46] | 0.26 [0.10,0.51] | 0.89 [0.43,1.74] |
| **Hispanic** | 1.54 [0.56,3.64] | 0.55 [0.20,1.30] | 0.16 [0.06,0.32] | 0.72 [0.33,1.47] |
| **Whites** | 1.71 [0.61,4.05] | 0.60 [0.22,1.42] | 0.22 [0.09,0.44] | 0.83 [0.39,1.66] |

In order to consider the approach to a risk limit of 3% cancer fatality [40] as recommended by the NCRP [1,13], we next performed calculations for lunar surface stays of 6-month as shown in **Table 7** for 45-year old females. In all cases average REID values for cancer risks are <3%, while the upper 95% confidence interval is 2.67% for API, 3.02% for Hispanic and White NS groups, and 3.27% for the NS Black group. These values are increased for NS by >25% when circulatory disease risks are included, and ∼15% for Average groups, however counting circulatory disease risks against limits falls outside of the previous NCRP recommendations [1,13]. **Figure 5** shows tissue specific risks predictions for Blacks and API females. In **Figure 6** the average values for these predictions are shown. The dominance of lung cancer risks is reduced to a great extent for NS. In contrast the increased lifespan shown in **Figures 1 and 2**, lead to slightly increased circulatory risks.

**Figure 5.**
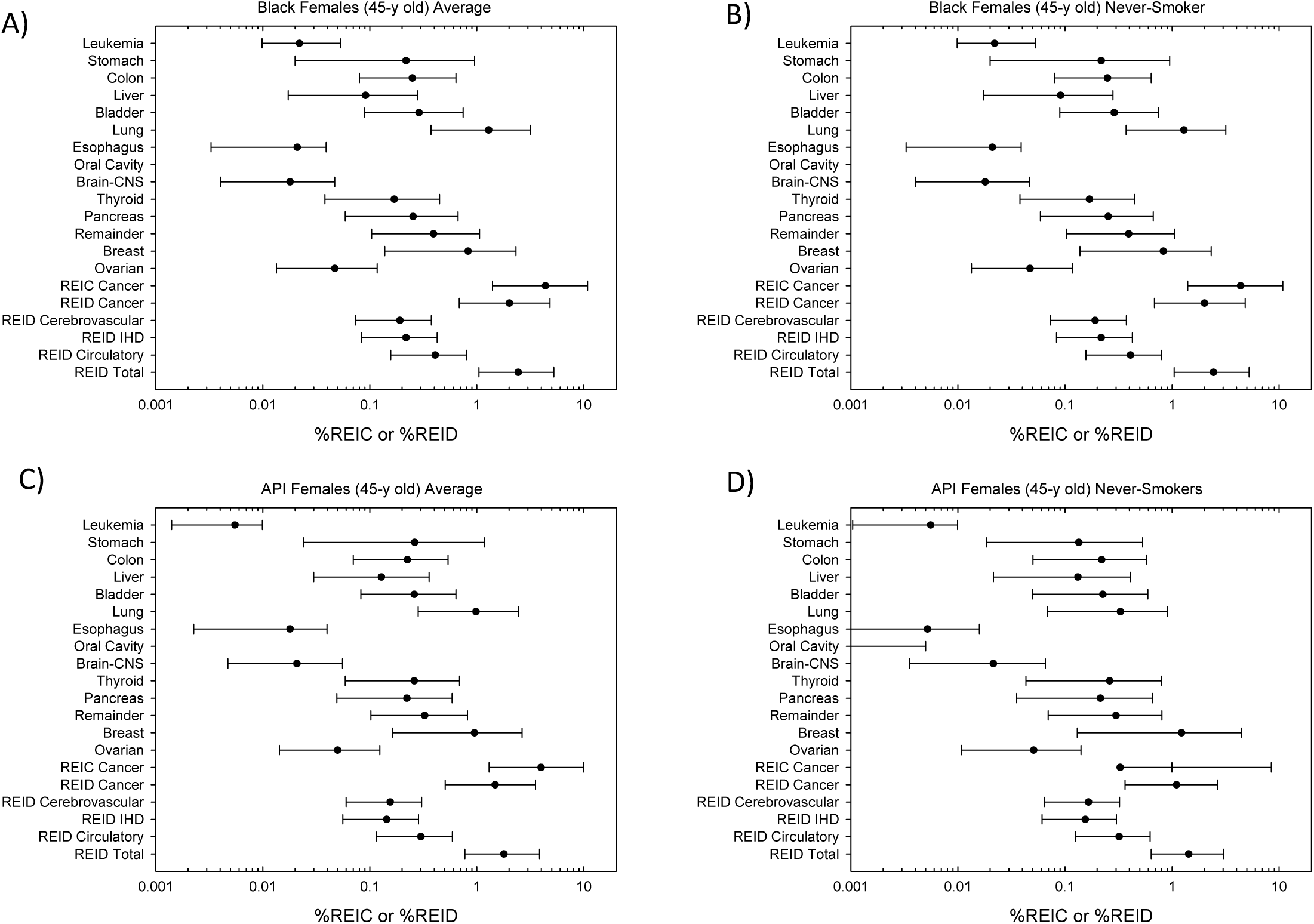
Tissue specific risks and 95% confidence intervals for Females on 80-day lunar missions. A) Black average population, B) Black never-smokers, C) Asian-Pacific Islander (API) average populations, and D) API never-smokers.

**Figure 6.**
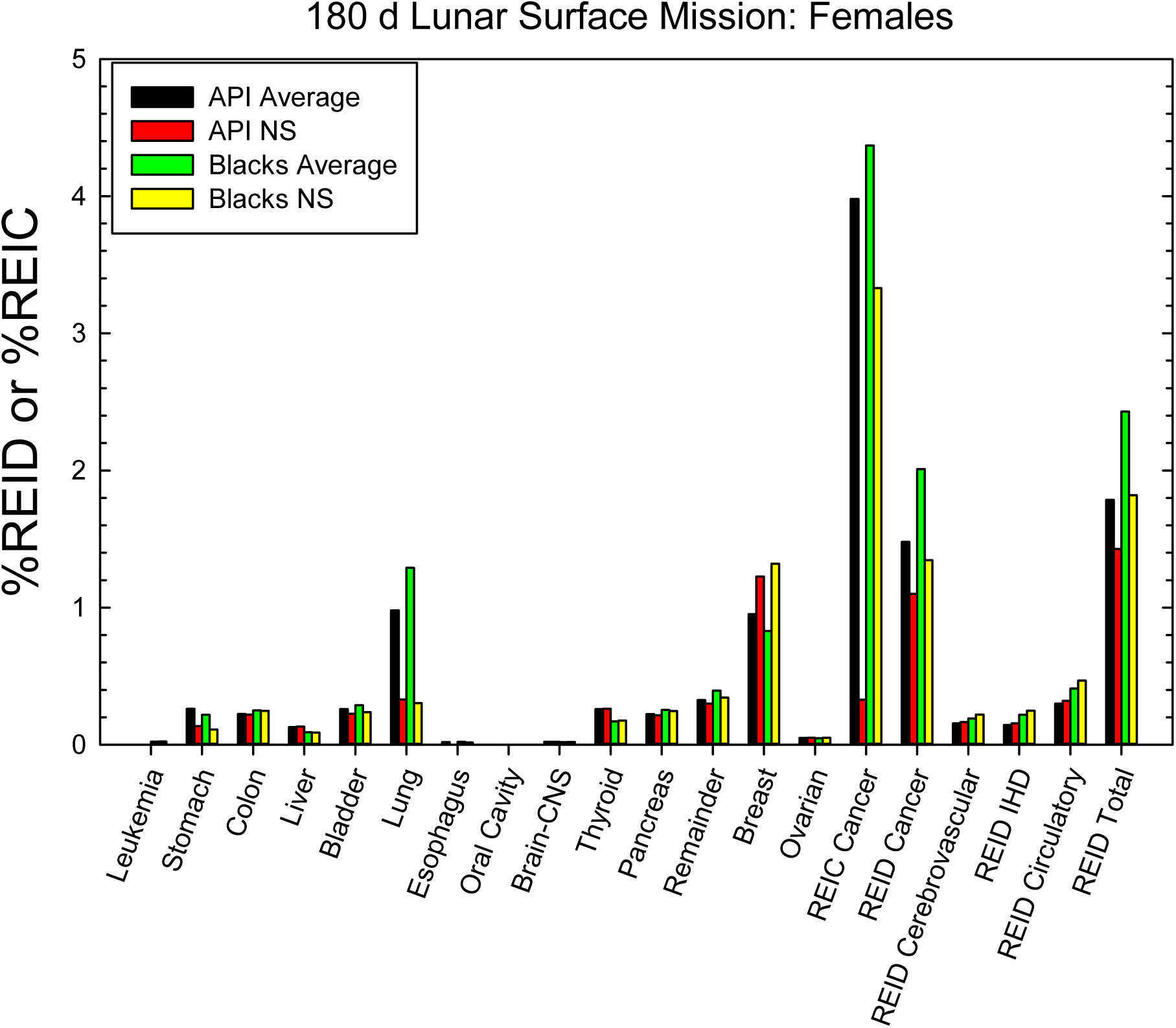
Average values for tissue specific cancers (%REIC) and %REID for cancer, circulatory diseases and total risk for API and Black females returning from 200-day lunar mission as described in text.

**Table 7.** Cancer and circulatory disease risks and 95% confidence intervals for 45-year old Female astronauts of several race or ethnic groups comparing never-smokers to average groups population in the USA. Lunar missions near solar minimum with 20 g/cm^2^ aluminum shield is considered in calculations. Mission total duration is 200 days with 20 days for Earth to Moon transit and return and 180 days on lunar surface.

| **Race-Ethnic Group** | **%REIC** | **%REID-cancer** | **%REID-circulatory** | **%REID-Total** |
| --- | --- | --- | --- | --- |
|  | **Average Population** |  |  |  |
| **API** | 3.98 [1.3,9.88] | 1.48 [0.51,3.53] | 0.30 [0.12,0.59] | 1.79 [0.77,3.85] |
| **Blacks** | 4.37 [1.4,10.8] | 2.01 [0.68,4.81] | 0.41 [0.16,0.80] | 2.43 [1.04,5.24] |
| **Hispanic** | 4.11 [1.35,10.1] | 1.45 [0.51,3.45] | 0.30 [0.11,0.58] | 1.75 [0.77,3.75] |
| **Whites** | 5.4 [1.7,13.4] | 2.2 [0.75,5.2] | 0.33 [0.13,0.64] | 2.52 [1.04,5.49] |
|  | **Never-Smokers** |  |  |  |
| **API** | 3.28 [1.0,8.46] | 1.10 [0.36,2.67] | 0.32 [0.13,0.63] | 1.43 [0.64,3.03] |
| **Blacks** | 3.33 [0.97,8.86] | 1.35 [0.44,3.29] | 0.47 [0.18,0.92] | 1.82 [0.82,3.81] |
| **Hispanic** | 3.69 [1.17,9.33] | 1.26 [0.43,3.02] | 0.31 [0.12,0.60] | 1.58 [0.69,3.37] |
| **Whites** | 3.64 [1.07,9.59] | 1.23 [0.40,3.02] | 0.38 [0.15,0.73] | 1.62 [0.71,3.43] |

## Discussion and Conclusions

In this report we extended previous research to model NS risks [8,9] and racial or ethnic group risks [12] into a combined model using the current version of NSCR denoted as NSCR-2022. Our results show important residual disparities in NS risk estimates, especially for Blacks with predictions of higher risks compared to other groups. We estimated uncertainties in risk estimates for each case considered, however the main focus of this report is to describe and consider the implications of various baseline factors for different population groups. The large uncertainties due to the sparsity of high LET and low dose radiobiology data leads to overlapping distributions for each group considered. However, differences in averages as large as 30% reported herein are similar to often studied variations in many other factors in space radiation protection, such as choice of shielding materials, large increases in shielding amounts, and extended GCR variations over the solar cycle.

The recent improved analysis of lifestyle factors in the analysis of LSS data by RERF [25-32], the CDC data on racial/ethnic and sex differences and the meta-analysis of data on S, FS, and NS mortality risks [6] suggest the differences considered in this report are important disparities for radiation risks that should be considered in risk assessment models and radiation protection. It is immature to such factors in the area of crew selection, however research in this area is needed. Of note is the large interaction between smoking and radiation risk for lung cancer in the LSS [25], while animal models used to assess RBE’s are independent of such interactions. The recent direct application of excess relative risk (DERR) model [41] suggested a much lower lung cancer risk for females than traditional models based on epidemiology data which are often confounded by primary and secondary smoke exposures of cohorts.

We used meta-analysis results for smoking related risks [6] mortality risks. These were applied to cancer incidence and fatality risk predictions. We considered tobacco related RR for the major causes of death of cancer, COPD, and circulatory diseases, however the inclusion of other less frequents causes of death such as digestive diseases and non-COPD respiratory diseases could increase our modeled life-tables for NS and therefore possibly increase their lifetime risk estimates from those made here. The study of Carter et al. [6] on smoking and mortality considered over 130,00 deaths to over 950,000 adults above age 55-y with data collected from 2000 to 2011. The percentage of persons using tobacco products has some age dependence with an increase in smoking cessation often occurring above age 70-y. For astronauts exposed in their 4^th^ or 5^th^ decade risks above age 50-y are the primary concern and this data set seems appropriate for our study. We did not consider racial or ethnic group dependences of RR for S, FS, or NS in our estimates. The cohorts in the meta-analysis of Carter et al. was ∼90% White and remainder Blacks. However, several studies [36,42-44] did not identify significant racial differences in *RR_S_* and *RR_FS_* for lung and other cancers. Future research for developing risk models for other groups such as FS is recommended by the authors.

## Data Availability

Data is included in manuscript or on request.

## Acknowledgements

No funding was received for this research.

